# Health literacy in the general population in the context of epidemic or pandemic coronavirus outbreak situations: rapid scoping review

**DOI:** 10.1101/2020.07.03.20145060

**Authors:** Uwe Matterne, Nina Egger, Jana Tempes, Christina Tischer, Jonas Lander, Marie-Luise Dierks, Eva-Maria Bitzer, Christian Apfelbacher

## Abstract

**BACKGROUND:** Authorities responded with contact restrictions and other measures to the global spread of SARS-CoV-2. Health literacy (HL) has been linked to health outcomes and refers to the ability to access, understand, appraise and apply health information in order to make good health decisions. When restrictions are gradually lifted, individual HL becomes essential to control the pandemic and to prevent the resumption of these restriction, should infection numbers surge again. The aim of this rapid scoping review, for which only studies from the general population were considered, was to describe the extent of existing research on HL in the context of previous coronavirus outbreaks (SARS-CoV-1, MERS-CoV and SARS-CoV-2). Facets of HL that were of particular interest were: type of assessment of HL (theory-based versus proxy assessment; validated instrument versus ad hoc assessment), domains of HL, interventions aiming to improve HL during outbreak situations, and HL surveillance during outbreak.

**METHODS:** We searched two major databases and included publications of quantitative and qualitative studies in English and German on any type of research on the functional, critical and communicative domains of HL conducted in the context of the three outbreaks in the general population. We extracted and tabulated relevant data and narratively reported where and when the study was conducted, the design and method used, and how HL was measured.

**FINDINGS:** 72 studies were included. Three investigated HL or explicitly referred to the concept of HL, 14 were guided by health behaviour theory. We did not find any study designed to develop or psychometrically evaluate pandemic HL instruments, or relate pandemic or general HL to a pandemic outcome, or any controlled intervention study. Type of assessment of the domains of HL varied widely.

**INTERPRETATION:** Theory-driven observational studies as well as interventions, examining whether pandemic-related HL can be improved are needed. In addition, the development and validation of instruments that measure pandemic-related HL is desirable.

## Introduction

In late 2019 an outbreak of a new viral disease occurred in Wuhan, China and later spread to almost all countries of the world ^1^. It is caused by a novel beta coronavirus, the Severe Acute Respiratory Syndrome – Coronavirus – 2 (SARS-CoV-2), which causes Coronavirus Disease (COVID) – 19)^2^. The clinical epidemiology of COVID-19 is currently being investigated intensely^3^. Course of disease may be very mild, asymptomatic to very severe with respiratory and systemic damage and requiring mechanical ventilation^4^. Responses of governments to the COVID-19 pandemic have been multi-faceted including outbreak management (suppression versus mitigation), provision of adequate clinical treatment facilities for severe cases and measures to alleviate the economic and psychosocial impact of the pandemic and the measures taken to manage it^2^. Public health measures implemented in many countries across the globe encompass contact restrictions and physical distancing, hygiene rules (i.e. frequent and thorough handwashing or disinfection), mask wearing, eye protection and recommendations about how to sneeze and cough ^3,5^. Some of these measures, particularly contact restrictions, have been law enforced in many countries ^6^. Relaxing regulations and re-organising social life requires people to voluntarily adhere to the named measures in order to avoid exponential growth of SARS-CoV-2 to reoccur. Further, people who contract SARS-CoV-2 need to know when and how to seek health care and/or be tested. Those who suffer from severe COVID-19 and survive will have to seek health care to mitigate the potentially long-lasting physical and psychological sequelae such as kidney damage^7^ or post-traumatic stress disorder^8^. In all these and other different scenarios, the concept of health literacy (HL) becomes a vital public health concept that is essential to counterpart on the individual level the social restrictions enforced by law. It “entails the motivation, knowledge and competencies to access, understand, appraise and apply health information in order to make judgements and take decisions in everyday life concerning healthcare, disease prevention and health promotion to maintain or improve quality of life throughout the course of life”^9^. When restrictions are gradually lifted, the role of individual level HL increases in order to prevent the resumption of these restriction, should infection numbers surge again.

In other words, what is necessary beyond governmental regulations and policy, is an increase in the levels of COVID-19 related health literacy^10,11^. We not only need to monitor the pandemic’s epidemiology during the course of the pandemic including the creation of herd immunity but also HL and health behavioural responses related to the pandemic in the population^11^. HL is considered a major determinant of a person’s health^12,13^, a factor that contributes to health inequalities^12^, and a person’s health behaviour, for instance, healthy diet adherence or non-smoking ^13^ and health care utilisation ^14^. There is evidence that lower HL is consistently associated with mortality ^14^ or lower self-rated health status^15^.

Research suggests that adequate HL may not be as prevalent among populations as might be necessary in order to navigate the increasingly complex healthcare landscape ^13,14,16^. Synthesised evidence suggests a relationship between levels of HL and infectious disease prevention in non-pandemic contexts. Inadequate HL was found to be associated with reduced adoption of protective behaviours such as vaccination uptake and poor understanding of antibiotics. Large research gaps were found in relation to infectious diseases with a high clinical and societal impact, such as tuberculosis and malaria^17^. For instance, it was emphasised that critical HL, which focuses on supporting effective political and social action, was not considered in any of the reviewed studies^17^. The strengths of this relationship may be exponentially higher under pandemic circumstances, but no synthesised information on this topic appears to exist to date. Further, the importance of individual HL in pandemic control has been emphasised more urgently^10,11^.

Therefore, the aim of this rapid scoping review, for which only studies from the general population were considered, was to describe the extent of existing research on HL in the context of previous coronavirus outbreaks (SARS-CoV-1, MERS-CoV and SARS-CoV-2). Facets of HL that were of particular interest were: type of assessment of HL (theory-based versus proxy assessment; validated instrument versus ad hoc assessment), interventions aiming to improve HL during outbreak situations, or HL surveillance during outbreak.

## Method

### Overview

This scoping review was performed according to the methodological framework as outlined by Khalil et al.^18^. Their guidelines regarding scoping reviews build on the work of Arksey and O’Malley’s five-stage scoping review framework^19^, complemented with the Joanna Briggs Institute methodology^20^, in order to (1) identify the research questions, (2) identify relevant studies, (3) select studies, (4) chart the data, and (5) collate and summarise the data. A scoping review’s objective is to identify the nature and extent of the existing evidence. Unlike other types of review, it does not endeavour to systematically evaluate the quality of available research, but rather seek to identify the contribution of existing literature to an area of interest ^21^. Our methodology was also guided by the rapid review approach which inevitable uses less rigor as necessary in a traditional systematic review due to the need for production within a short time-frame using limited resources^22^. The protocol for this rapid review was registered at OSFREGISTRIES on 06/04/2020^23^.

### Search strategy, selection criteria, extraction strategy and data analysis

Two authors (UM, NE) ran the search strategy on PubMed (MEDLINE®) and PsycINFO® on 20^th^ April 2020. Citations were downloaded to Citavi (Swiss Academic Software). We included publications in English and German of quantitative and qualitative studies. The same authors evaluated titles and abstracts excluding any irrelevant ones. Full texts of the remaining citations were obtained, and two authors (UM, NE) reviewed these, excluding any, which did not meet the inclusion criteria. Finally, reference lists of remaining papers were hand-searched for additional relevant studies. We then compared results from full text screening; there were only minor discrepancies, which were resolved through discussion with the whole team. Data extraction was carried out by five authors (UM, NE, JT, CT, JL) in independent pairs of two. Consensus was achieved through discussion and arbitration within the team. The search strategy was informed by HL theory (derivation of search terms) and is displayed in appendix **1**. Inclusion criteria were:

We included reports on any type of research on the functional, critical and communicative domains of HL^24^ conducted in the context of SARS-CoV-2, SARS-CoV-1 and MERS-CoV in the general population. This was a rational decision as an initial search using HL as the chief search term in conjunction with the aforementioned coronavirus outbreaks resulted in very few hits. We used the following definitions / concepts of functional, communicative and critical HL: Functional HL is broadly compatible with the narrow definition of ‘health literacy’ which can be considered to consist of health-related knowledge, attitudes, motivation, behavioural intentions, personal skills, or self-efficacy^24^. Communicative HL means to be able …’to derive meaning from different forms of communication’…, while the ability to critically analyse information is referred to as critical HL^24^.

The following data were extracted from the included studies: authors, publication year, country of study, type of epidemic or pandemic outbreak (SARS-CoV-2, SARS-CoV-1, MERS-CoV), participants (including sample size), design, method, and instruments, and measured constructs including how they were measured (only if applicable e.g. not in qualitative studies). Findings were synthesized quantitatively and narratively and reporting followed the guidelines as proposed in PRISMA-ScR^25^. A critical appraisal of the quality of the included studies was not within the scope of this review. We do however, comment on major methodological issues regarding the studies.

There was no funding source for this study.

## Results

The search in PubMed (MEDLINE®) and PsycInfo® yielded 3394 references, two were obtained from colleagues^26,27^, leading to 2766 references after removal of duplicates. Title and abstract screening resulted in exclusion of 2652 articles. Full-texts of the remaining 114 references were assessed for eligibility leading to inclusion of 77 publications pertaining to 72 studies. Details of the selection stages are provided in Figure **1**. SARS-CoV-2 was investigated in 10, MERS-CoV in 26 and SARS-CoV-1 in 36 studies. Only three studies investigated HL or explicitly referred to the concept of HL^27–29^. 14 studies were conducted in the context of health behaviour theory, seven of another theory (Appendix **2**). All studies, while mainly not explicitly investigating HL, measured one or more components of HL (Appendix **2**). Most studies were observational or short longitudinal (58 cross-sectional, eight pre-post) and six qualitative. All SARS-CoV-two studies were conducted during, of the MERS-CoV studies 27 during, one during (first wave), eight after, of the MERS-CoV studies 24 during, one after and one both during and after the pandemic/outbreak. 66 studies used questionnaires, two focus group discussion, four other qualitative methods (e.g. interviews) for data collection. 49 studies studied convenience or opportunity samples and 23 representative samples drawn from general populations. Sample size ranged from 19 – 222.599 participants.

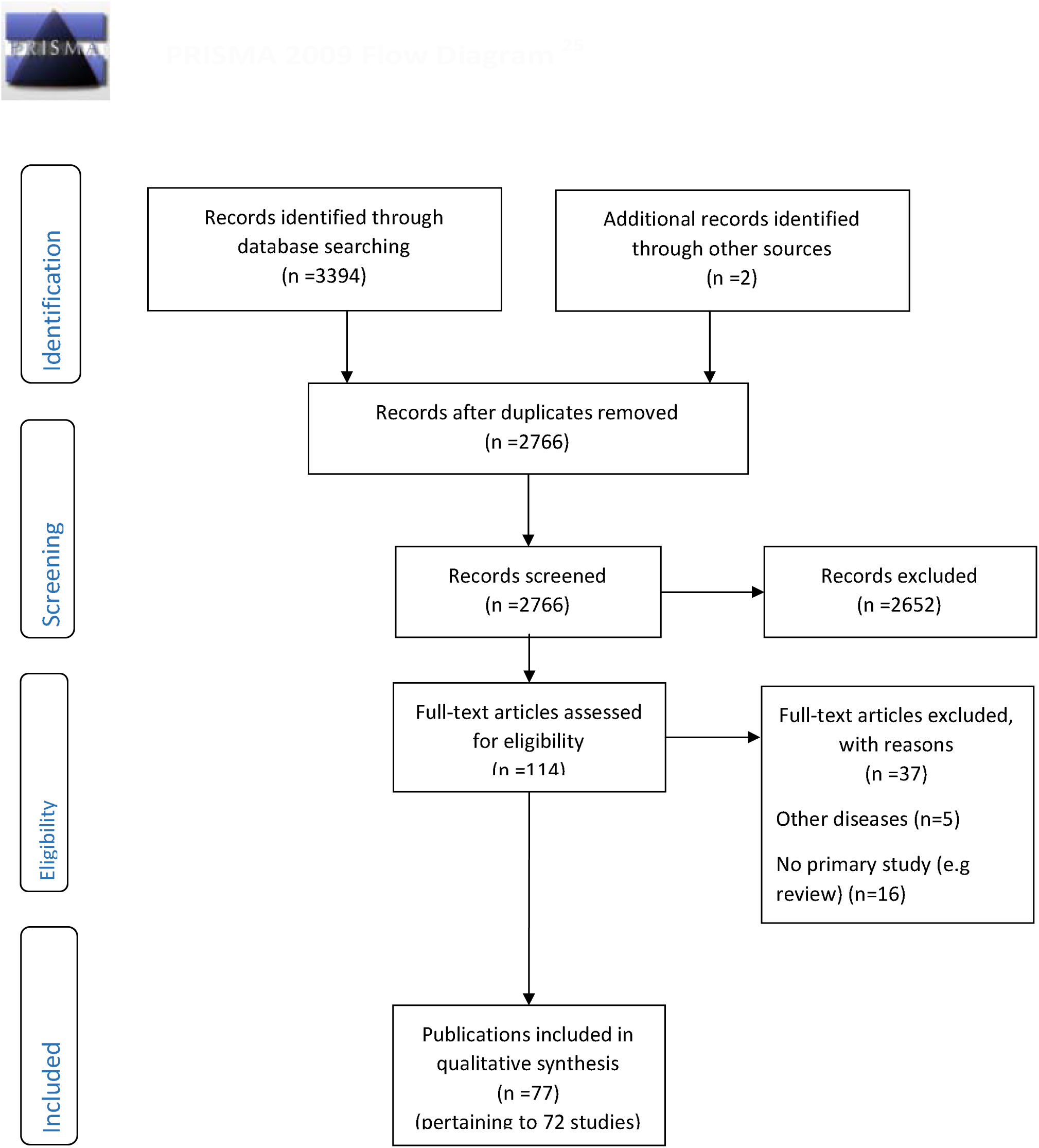
PRISMA 2009 Flow Diagram ^25^.

### Functional, critical, communicative HL and individual behaviour in quantitative studies

Within the nine quantitative SARS-CoV-2 studies knowledge was measured in seven, attitude in seven, risk perceptions in four, SE in three, critical HL in five, communicative HL in three, Health-information seeking behaviour (HISB) in two, and behavioural aspects in four studies. Only one study ^26^ measured functional, critical and communicative HL as well as a behavioural outcome. All others assessed only some aspects of HL. While seven studies reported on knowledge, most studies asked only about knowledge of symptoms, no study undertook a comprehensive assessment covering a broad range of SARS-CoV-2 related knowledge. Within the knowledge domain, symptoms were most often assessed. (Table **1**). Wearing a mask was the most frequently assessed behaviour (Table **1**).

**Table 1:** Health literacy (HL) and behavioural dimensions assessment in quantitative studies.

|  |  | SARS-CoV-2<br>9 studies | SARS-CoV-1<br>31 studies | MERS-CoV<br>26 studies |
| --- | --- | --- | --- | --- |
| Functional HL | Knowledge dimension | 7 26,29,33,34,56-60 | 25 30,61-66,68-74,76-80,82-86,89,90,92 | 21 31,35,93-106,109-114 |
|  | Awareness | 2 58,59 | 3 64,83,89 | 9 97,98,100,101,104-106,109,110,113 |
|  | Nature of disease | 2 26,59 | 10 30,61-64,66,71,85,89,92 | 9 35,95,96,100,102,105,110,111,114 |
|  | Transmission mode | 4 26,33,34,58,60 | 18 61-63,65,66,70-74,76-80,82-85 | 18 31,35,93-96,99-102,104-106,109-111,113,114 |
|  | Symptoms | 6 26,29,56-60 | 8 30,61,62,66,70,85,89,92 | 13 31,35,93-100,102,110,111,114 |
|  | Course | 2 26,29 | 4 66,73,74,89 | 7 35,94-96,101,111,114 |
|  | Treatment | 2 26,33,34 | 5 62,63,70,74,89 | 8 31,35,94,95,100,109,111,114 |
|  | Risk groups | 1 56 | 0 | 3 35,94,100 |
|  | Fatality rate/# infections | 2 33,34,56,57 | 1 64 | 3 35,103,111 |
|  | Measures of infection control | 0 | 1 83 | 1 100 |
|  | Prevention | 4 29,56,57,59,60 | 3 70,71,74 | 8 35,93,94,96,101,102,110,114 |
|  | Attitude | 7 26,33,34,56-60 | 28 30,61-63,65-82,84,86-92 | 17 31,32,35,93-99,101-105,108,112,114 |
|  | Risk perception | 4 26,33,34,56,57 | 20 30,64,65,70-79,81,82,84,86,87,89,90,92 | 18 31,32,35,93-102,104,105,110,111,114,116 |
|  | Self-efficacy/skills/preparedness | 3 26,27,29 | 8 30,64,66,81,86,87,90,92 | 3 31,32,116 |
| Critical HL |  | 5 26,27,29,33,34,60 | 11 30,61,64,70-72,81,82,86,89,92 | 4 32,108,113,114 |
| Communicative HL |  | 3 26,27,58 | 12 30,61-65,71-73,81,83,89 | 17 31,93,96-100,102-105,107,109-111,113,115,116 |
| HISB |  | 2 26,29 | 4 61,63,66,89 | 2 31,107 |
| Behaviour/practices |  | 4 26,29,33,34,60 | 18 61,63-66,71,73,75-79,82,84-87,89,90 | 10 31,35,95,97-99,106-108,114,115 |
|  | Wash hands | 1 33,34 | 13 64-66,73,76-79,82,84,85,87,89,90 | 9 31,35,95,97-99,106-108,115 |
|  | Wear mask | 3 26,33,34,60 | 15 64-66,73,75-79,82,84-87,89,90 | 7 31,35,95,97-99,106,108 |
|  | Avoid touching face | 0 | 3 76-79 | 4 35,95,99,108 |
|  | Cough/Sneeze hygiene | 1 33,34 | 7 65,76-79,82,84,90 | 6 31,35,95,97-99,108 |
|  | Correct tissue disposal | 0 | 0 | 3 35,95,97,98 |
|  | Self-impose quarantine if experiencing symptoms | 0 | 1 75 | 2 35,95 |
|  | Consult doctor if experiencing symptoms | 0 | 5 63,66,75,87,89 | 1 35 |
|  | Avoid sharing personal items | 1 33,34 | 6 65,76-79,82,84 | 0 |
|  | Avoid travelling | 0 | 3 61,63,89 | 0 |
|  | Avoid travelling to high-risk areas | 0 | 2 71,89 | 0 |
|  | Avoid public transport | 0 | 4 64,73,75,89 | 0 |
|  | Avoid crowded places | 2 26,60 | 5 63,64,71,73,89 | 1 108 |
|  | Avoid eating out | 0 | 3 64,66,89 | 0 |
|  | Avoid people with flu-like symptoms | 0 | 2 61,66 | 2 99,108 |
|  | General personal self-care (pro-active HB) | 0 | 6 64,66,85,87,89,90 | 0 |
|  | Home hygiene | 0 | 7 63,64,66,73,85,87,90 | 1 99 |
|  | Avoid camel exposure and products | 0 | 0 | 3 97-99,106 |
|  | Other behavioural responses | 2 26,29 | 7 63,65,66,73,84,85,89 | 3 106-108 |

HB: health behaviour; HISB: health-information seeking behaviour; note: cited references do not correspond to number of studies but publications

31 quantitative studies were conducted in the context of SARS-CoV-1. 25 measured knowledge, 28 attitude, 20 risk perception, eight SE, 11 critical HL, 12 communicative HL, and 18 behaviour. One study^30^ reported all six HL aspects, the others one to five aspects. Within knowledge, transmission mode was most often measured. Although 25 studies reported knowledge assessment, most studies did not comprehensively assess knowledge (Table **1**). Handwashing was the most frequently measured behaviour (Table **1**).

Of the 26 quantitative MERS-CoV studies 21 measured knowledge, 17 attitude, 18 risk perceptions, 3 SE, 4 critical HL, 17 communicative HL, and 10 behaviour. Two studies assessed five of the six HL aspects ^31,32^, the remainder one to four. Within knowledge, transmission mode was most often assessed. Again, most studies did not comprehensively assess knowledge. Handwashing was the most frequently assessed behaviour (Table **1**).

The reported measured depth within the domains of HL varied widely among the studies (results not shown). For instance, the number of knowledge components ranged from one to at least eight.

### Health-information seeking behaviour (HISB) in quantitative studies

HISB was measured in two (SARS-CoV-2), four (SARS-CoV-1), two (MERS-CoV) quantitative studies.

### Pandemic HL measurement and relationship to pandemic outcome (quantitative and qualitative studies)

Our search failed to come across any studies designed to develop or psychometrically evaluate pandemic HL instruments or relate pandemic or general HL to a pandemic outcome.

### Other aspects of HL measurement in pandemic contexts (quantitative and qualitative studies)

The number of items per HL aspect varied widely (data not shown), hardly any study reported on psychometric properties, two studies from three publications^28,33,34^ were the notable exception (Appendix 3) and a clear distinction between knowledge, attitude, or risk perceptions was sometimes absent. For instance, perceived vulnerability was reported as an attitude^35^.

### Qualitative studies

Six studies explored domains of HL in the context of SARS-CoV-2, SARS-CoV-1, and MERS-CoV. One study^36^ reported low risk perceptions and a lack of seeking relevant health information in relation to SARS-CoV-2. Two studies^37,38^ explored risk perceptions and preventive behaviour in relation to SARS-CoV-1, another^39^ explored individual experiences during quarantine. One study reported low knowledge about SARS-CoV-1 and its prevention^40^. Another study in the context of SARS-CoV-1 concluded that attitudes towards mask wearing had substantially changed in the post-SARS-CoV-1 period^41^.

## Discussion

While individual HL is recognised as an increasingly important construct in public health^42^, it is of note that only three studies emerged from our extensive search, which explicitly referred to the construct of HL in the context of any of the three coronavirus outbreaks. One used the Newest Vital Sign (NVS), a test measuring nutrition label information processing ability^29^, another study^28^ administered a short form of the HLS-EU-Q47, an HL instrument rooted in testable theory^43^ and the third ^27^ study used a version of the HLS-EU-Q47 adapted to SARS-CoV-2. However, the latter provided no evidence on the psychometric properties of the adapted instrument. Hence, at present there seems to be no tested instrument designed to measure coronavirus pandemic-related HL. There is, however, one HL instrument assessing print and multimedia literacy in respect to respiratory diseases^44^.

Most of the other included studies were not theory-based. It is important to highlight that these studies did not purport to measure HL, but were included in this review because the search strategy was based on a pragmatic application of suggested HL components within domains ^24,45^. Of those that were theory-driven, the majority employed health-behaviour theory as conceptualised by social cognition models. There is substantial overlap between socio-cognitive predictors of health behaviour and HL. For instance, attitude and self-efficacy (defined as behavioural control) are part of the theory of planned behaviour^46^, risk perceptions part of the health belief model^47^ or knowledge part of protection motivation theory^48^. Theory-based research allows the formulation of testable a priori hypotheses, and if necessary revision of the theory. Nonetheless, the measures obtained from those studies lacking an explicit theoretical foundation can be considered proxies of HL because they constitute or at least contribute to one or more HL domains.

While there appears to be no evidence linking validly measured (epidemic or pandemic) HL to coronavirus outbreak/pandemic outcomes there is evidence that HL can be linked to other epidemic outbreaks, e.g. the 2014–2016 Ebola epidemic outbreak in West Africa resulted among other factors from low health literacy^49^. A Center for Disease Control and Prevention campaign, with input from partners, helped increase HL^50^. HL has also been shown to be associated with health and health behaviour in general. Hence, one would expect that this association would hold in coronavirus outbreak situations.

Communicative HL included the measurement of access to different sources of information. Whether this had anything to do with better decisions about health in relation to any of the three outbreaks, remained unclear. Knowledge items were generally devised by the authors, and very few reported to have items checked against guidelines. This and the lack of an objective standard for cut-offs make knowledge assessment arbitrary as it cannot be established whether knowledge items reflect current and correct evidenced knowledge. Similarly, while risk perceptions generally pertain to perceptions of vulnerability/susceptibility to and severity of a disease, they were not always measured accordingly or sometimes subsumed under the term attitudes or knowledge.

We also observed very little evidence about the psychometric properties of instruments used to measure the socio-cognitive variables attitude, risk perceptions, and self-efficacy. It is desirable to know whether measures are reliable and valid, and sensitive to change if the aim is to reflect the effects of health literacy interventions by e.g. education (responsiveness).

Even if knowledge, attitudinal constructs, risk perceptions or self-efficacy were composed in a clear-cut and unequivocal way and psychometrically sound, uncertainty as to whether HL in its broader definition^9,51^ as a composite/compound construct was measured, would still prevail. HL was proposed to be a latent construct^43^ thus indicators for its measurement are necessary. There is the need for the development of adequate measurement models.

The present review cannot ascertain, whether established instruments such as TOFHLA (Test of Functional Health Literacy)^53^, or the broader dimension based instruments, for instance the HLQ (Health Literacy Questionnaire)^54^ could be used to predict a pattern of association between HL and epidemic or pandemic outcomes (and antecedents such as favourable behaviours and practices), because no such investigations appear to have been carried out, yet. The study^28^ that used a short form of the HLS-EU-Q did not investigate the relationship between HL and pandemic outcome/preventive behaviour but coping responses to the outbreak (depression, quality of life). Okan et al.^27^ reported individuals’ subjective perceptions about how well they could access, understand, appraise and apply information in the SARS-CoV-2 context but did not test the actual level of what these skills pertain to and whether they are related to better/more favourable behaviour/practices.

Further, it also not possible to state at present whether pandemic outbreaks require a specific HL instrument, that is able to explain variance in relevant behaviour and practices over and above that of general instruments (i.e. latent trait/construct measured by discrete manifest cognitive antecedents of behaviour).

In this rapid review, it was not possible to adhere to the methodological rigour that is expected from a standard scoping review. As this review was conducted as a scoping review, we did not look at the strengths of any reported associations between HL aspects and behavioural aspects. Further, it is beyond the scope of this review to assess the quality of the reviewed studies according to standard guidelines for observational studies.

At present HL in the context of coronavirus outbreaks is at an early stage to inform public health/educational strategies aimed at improving the public’s HL in order to contain the spread of pandemics. One study^26^ appears to be able to shed light on the question whether HL related aspects change over the course of the pandemic as its survey is conducted in weekly intervals.

We recommend future research be guided theory from HL research ^9,45^ in the much needed work on HL in pandemic outbreak situations. Consequently, assessment of HL should be based on the ability to access, understand, critically appraise and eventually apply information to make better choices about one’s health in pandemic outbreak situations when viewed as a set of meta-cognitive skills or a latent trait^43^. Nevertheless, operationalisations at the manifest level, for example, knowledge, or attitudes (which influence critical appraisal) need to be considered, as latent constructs cannot be directly measured. Nevertheless, in the interim, public health communication could benefit from what is generally known from HL research. Health information should be clear so that all members of the public can access needed health information for routine and critical decisions^55^.

Beside theory-driven observational studies, we also need interventions, examining whether coronavirus pandemic-related HL can be improved. In addition, research should also attempt to develop HL instruments that measure coronavirus pandemic-related HL and test the reliability, validity and responsiveness to change. The latter is of particular importance, if we want to be able to examine change during the stages of a pandemic.

## Data Availability

n/a

## Contributors

CA conceived the study. CA & UM developed the protocol. UM, NE, CT, JT, JL, CA & EMB contributed to the search strategy, screened the search results, extracted data, and interpreted the findings. UM, CA & EMB wrote the paper. All authors reviewed the paper for important intellectual content.

## Acknowledgments

There was no funding source for this study.

## Appendix 1: search strategy

PubMed (MEDLINE^®^)

(health literacy[MeSH Terms] OR (health[Title/Abstract] AND competence[Title/Abstract]) OR literacy[Title/Abstract] OR knowledge[Title/Abstract] OR attitude[Title/Abstract] OR motivation* OR intention* OR skills OR self-efficacy[MeSH] OR organisation*[Title/Abstract] OR community[Title/Abstract]) AND ((2019-nCoV OR 2019nCoV OR COVID-19 OR SARS-CoV-2 OR ((wuhan AND coronavirus) AND 2019/12[PDAT]:2030[PDAT])) OR (“Severe Acute Respiratory Syndrome”[MeSH Terms] OR SARS) OR (“middle east respiratory syndrome coronavirus”[MeSH Terms] OR MERS)) NOT (animals [mh] NOT humans [mh])

PsycINFO^®^

(health literacy or health education or health knowledge or health information or health understanding) AND (pandemic* or epidemic* or outbreak or covid-19 or coronavirus OR 2019-ncov or sars OR sars-cov-2 OR mers) NOT (hiv or aids or acquired human immunodeficiency syndrome or human immunodeficiency virus)

## Appendix 2: Summary of study characteristics

| Study ID | Country | n | When conducted | Design | Method | Sampling method | HL specific | Other theory guided | Type of theory |
| --- | --- | --- | --- | --- | --- | --- | --- | --- | --- |
| **SARS-CoV-2** |  |  |  |  |  |  |  |  |  |
| Betsch 2020<sup>26</sup> | GER | 1000 | during | obs cross | Quest. | repr. | no | no | n.a. |
| Geldsetzer 2020a 2020b<sup>56,57</sup> | UK, USA | 5974 | during | obs cross | Quest. | Conv. | no | no | n.a. |
| Khan 2020<sup>58</sup> | PAK | 302 | during | obs cross | Quest. | Conv. | no | no | n.a. |
| Ma 2020<sup>36</sup> | AUS | 28 | during | qual. | Focus groups | Conv. | no | no | n.a. |
| Nguyen 2020<sup>28</sup> | VNM | 3974 | during | obs cross | Quest. | Conv. | yes | no | n.a. |
| Okan 2020<sup>27</sup> | GER | 1000 | during | obs cross | Quest. | repr. | yes | no | n.a. |
| Roy 2020<sup>59</sup> | IND | 662 | during | obs cross | Quest. | Conv. | no | no | n.a. |
| Wang 2020a 2020b<sup>33,34</sup> | CHN | 1738 | during | obs Pp | Quest. | Conv. | no | no | n.a. |
| Wolf 2020<sup>29</sup> | USA | 630 | during | obs cross | Quest. | Conv. | yes | no | n.a. |
| Zhong 2020<sup>60</sup> | CHN | 6919 | during | obs cross | Quest. | Conv. | no | no | n.a. |
| **SARS-CoV-1** |  |  |  |  |  |  |  |  |  |
| Bener 2004<sup>61</sup> | QAT | 1386 | during | obs cross | Quest. | Conv. | no | no | n.a. |
| Bergeron 2005<sup>62</sup> | CAN | 300 | post | obs cross | Quest. | Conv. | no | no | n.a. |
| Blendon 2004<sup>63</sup> | CAN, USA | 1500 | during | obs cross | Quest. | repr. | no | no | n.a. |
| Brug 2004<sup>64</sup> | NLD | 373 | during | obs cross | Quest. | unclear | no | no | n.a. |
| Cava 2005<sup>39</sup> | CAN | 21 | during | qual. | Interview | Conv. | no | no | n.a. |
| Chan 2007<sup>65</sup> | HKG | 296/122 | during | obs Pp Intervention | Quest. | Conv. | no | no | n.a. |
| Cheng 2006<sup>66</sup> | CHN, SGP, HKG, CAN | 300 | during | obs Pp | Quest. | Conv. | no | yes | THB |
| Chuo 2014<sup>67</sup> | TWN | 294/88 | partly during (first survey wave) | obs Pp | Quest. | Conv. | no | yes | THB |
| Des Jarlais 2006/2005<sup>68,69</sup> | USA | 928 | during | obs cross | Quest. | repr. | no | yes | OM |
| Deurenberg-Yap 2005<sup>70</sup> | SGP | 853 | during | obs cross | Quest. | repr. | no | yes | OM |
| Hazreen 2005<sup>71</sup> | MYS | 201 house holds | during | obs cross | Quest. | repr. | no | no | n.a. |
| Ishizaki 2004<sup>72</sup> | JPN | 821 | during | obs cross | Quest. | Conv. | no | yes | THB |
| Jiang 2009<sup>38</sup> | UK, NLD | 164 | post | Qual. | Focus groups | Conv. | no | yes | GT |
| Lau 2003<sup>73</sup> | HKG | 1397 | during | obs Pp | Quest. | repr. | no | yes | THB |
| Lau 2005<sup>74</sup> | HKG | 820 | during | obs cross | Quest. | repr. | no | no | n.a. |
| Lau 2004<sup>75</sup> | HKG | 820 | during | obs cross | Quest. | Conv. | no | yes | THB |
| Leung 2003<sup>76</sup> | HKG | 1115 | during | obs cross | Quest. | repr. | no | no | n.a. |
| Leung 2005<sup>77</sup> | HKG | 4481 | post | obs Pp | Quest. | repr. | no | no | n.a. |
| Leung 2004/2009<sup>78,79</sup> | HKG, SGP | 1906 | during | obs cross | Quest. | repr. | no | no | n.a. |
| Lim 2003<sup>80</sup> | SGP | 101 | during | obs cross | Quest. | Conv. | no | no | n.a. |
| Peng 2010<sup>81</sup> | TWN | 1278 | post | obs cross | Quest. | repr. | no | no | n.a. |
| Quah 2004<sup>82</sup> | SGP | 1201 | during | obs cross | Quest. | repr. | no | no | n.a. |
| Seng 2004<sup>83</sup> | SGP | 593 | post | obs cross | Quest. | Conv. | no | no | n.a. |
| Siu 2016<sup>41</sup> | HKG | 40 | post | qual. | interview | Conv. | no | no | n.a. |
| So 2004<sup>84</sup> | HKG | 163/112 | during | obs cross | Quest. | Conv. | no | no | n.a. |
| Tan 2004<sup>85</sup> | CHN | 1807 | during | obs retros | Quest. | Conv. | no | no | n.a. |
| Tang 2003<sup>86</sup> | HKG | 2331 | during | obs cross | Quest. | Conv. | no | yes | THB |
| Tang 2005<sup>87</sup> | HKG | 354 | during | obs cross | Quest. | Conv. | no | yes | THB |
| Tracy 2009<sup>88</sup> | CHN | 500 | post | obs cross | Quest. | Conv. | no | yes | PHM |
| Tse 2003<sup>40</sup> | HKG | 40 | during | qual. | interview | Conv. | no | no | n.a. |
| Vartti 2009<sup>89</sup> | NLD, FIN | 681 | during | obs cross | Quest. | Conv. | no | yes | MISR |
| Voeten 2009<sup>30</sup> | UK, NLD | 300 | post | obs cross | Quest. | Conv. | no | yes | THB |
| Wills 2008<sup>37</sup> | CAN | 19 | during | qual. | interview | Conv. | no | no | n.a. |
| Wong 2005<sup>90</sup> | HKG | 230 | during | obs cross | Quest. | repr. | no | yes | THB |
| Yip 2007<sup>91</sup> | HKG | 463 | during | obs cross | Quest. | Conv. | no | no | n.a. |
| Zwart 2009<sup>92</sup> | DNK,<br>NLD,<br>UK,<br>ESP,<br>POL,<br>SGP,<br>CHN,<br>HKG | 3436 | during | obs cross | Quest. | repr. | no | yes | THB |
| **MERS-CoV** |  |  |  |  |  |  |  |  |  |
| Al-Hazmi 2018<sup>93</sup> | SAU | 1109 | during | obs cross | Quest. | Conv. | no | no | n.a. |
| Al-Mohaissen 2017<sup>94</sup> | SAU | 1541 | during | obs cross | Quest. | Conv. | no | no | n.a. |
| Alhomoud 2017<sup>35</sup> | SAU | 292/25<br>7 | during | obs cross | Quest. | Conv. | no | no | n.a. |
| Almutairi 2015<sup>95</sup> | SAU | 1147 | during | obs cross | Quest. | Conv. | no | no | n.a. |
| Alotaibi 2017<sup>96</sup> | SAU | 417 | during | obs cross | Quest. | Conv. | no | no | n.a. |
| Alqahtani 2016a/2016b<sup>97,98</sup> | AUS | 421 | during | obs Pp | Quest. | Conv. | no | yes | THB |
| Alqahtani 2017<sup>99</sup> | SAU,<br>KWT,<br>UAE,<br>QAT,<br>BHR,<br>OM | 1812 | during | obs cross | Quest. | Conv. | no | yes | THB |
| Althobaity 2017<sup>100</sup> | SAU | 2120 | during | obs cross | Quest. | Conv. | no | no | n.a. |
| Ashok 2016<sup>101</sup> | SAU | 404 | during | obs cross | Quest. | Conv. | no | no | n.a. |
| Bawazir 2018<sup>102</sup> | SAU | 676 | during | obs cross | Quest. | Conv. | no | no | n.a. |
| Gautret 2013<sup>103</sup> | FRA | 360 | during | obs cross | Quest. | Conv. | no | no | n.a. |
| Hoda 2016<sup>104</sup> | SAU | 854/65<br>8 | during | obs cross | Quest. | repr. | no | yes | CPM |
| Hou 2018<sup>105</sup> | SGP | 2969 | during | obs cross | Quest. | Conv. | no | yes | THB |
| Jang 2018<sup>31</sup> | KOR | 1036 | during | obs cross | Quest. | repr. | no | yes | THB |
| Kamau 2019<sup>106</sup> | KEN | 22 | during | obs cross | Quest. | Conv. | no | no | n.a. |
| Kim 2018<sup>32</sup> | KOR | 814 | during | obs cross | Quest. | Conv. | no | yes | VBM |
| Lee 2019<sup>107</sup> | KOR | 855 | post | obs cross | Quest. | repr. | no | no | n.a. |
| Lee 2016<sup>108</sup> | KOR | 6739 | during | obs cross | Quest. | repr. | no | no | n.a. |
| Lin 2017<sup>109</sup> | USA | 627 | both | obs cross | Quest. | repr. | no | no | n.a. |
| Migault 2019<sup>110</sup> | FRA | 82 | during | obs Pp<br>Intervention | Quest. | Conv. | no | no | n.a. |
| Nooh 2010<sup>111</sup> | SAU | 384 | during | obs cross | Quest. | repr. | no | no | n.a. |
| Sahin 2015<sup>112</sup> | TUR | 381 | during | obs cross | Quest. | Conv. | no | no | n.a. |
| Tashani 2014<sup>113</sup> | AUS | 119 | during | obs cross | Quest. | Conv. | no | no | n.a. |
| Yang 2017<sup>114</sup> | KOR | 1470 | during | obs cross | Quest. | Conv. | no | no | n.a. |
| Yang 2019<sup>115</sup> | KOR | 222,59<br>9 | during | obs cross | Quest. | repr. | no | no | n.a. |
| Yoo 2016<sup>116</sup> | KOR | 1000 | during | obs cross | Quest. | repr. | no | no | n.a. |

CPM: communication-persuasion model; Conv.: convenience or opportunity sample; GT: grounded theory; HL specific: Do the authors specifically refer to health literacy and/or a health literacy concept?; MISR: Model of Illness Perceptions and Self-Regulation; n.a.: not applicable; obs cross: observational cross-sectional; obs Pp: observational pre-post; OM: other model; THB: Theory of health behaviour; PHM: Public health model; repr.: representative; VBM: value-based model; Quest.: Questionnaire

## Appendix 3: Health literacy (HL) dimensions and their components and behaviour/practice measured in studies

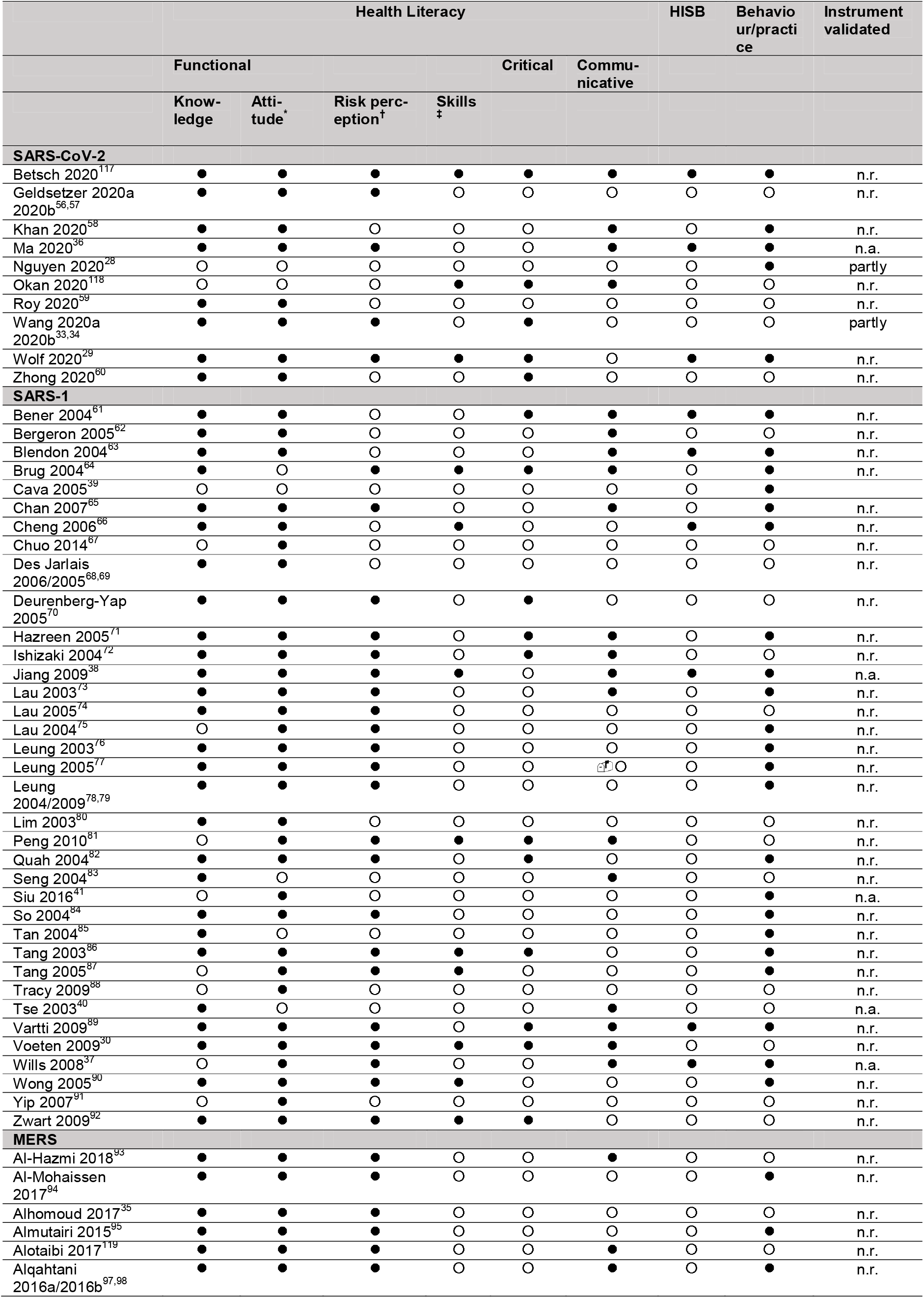
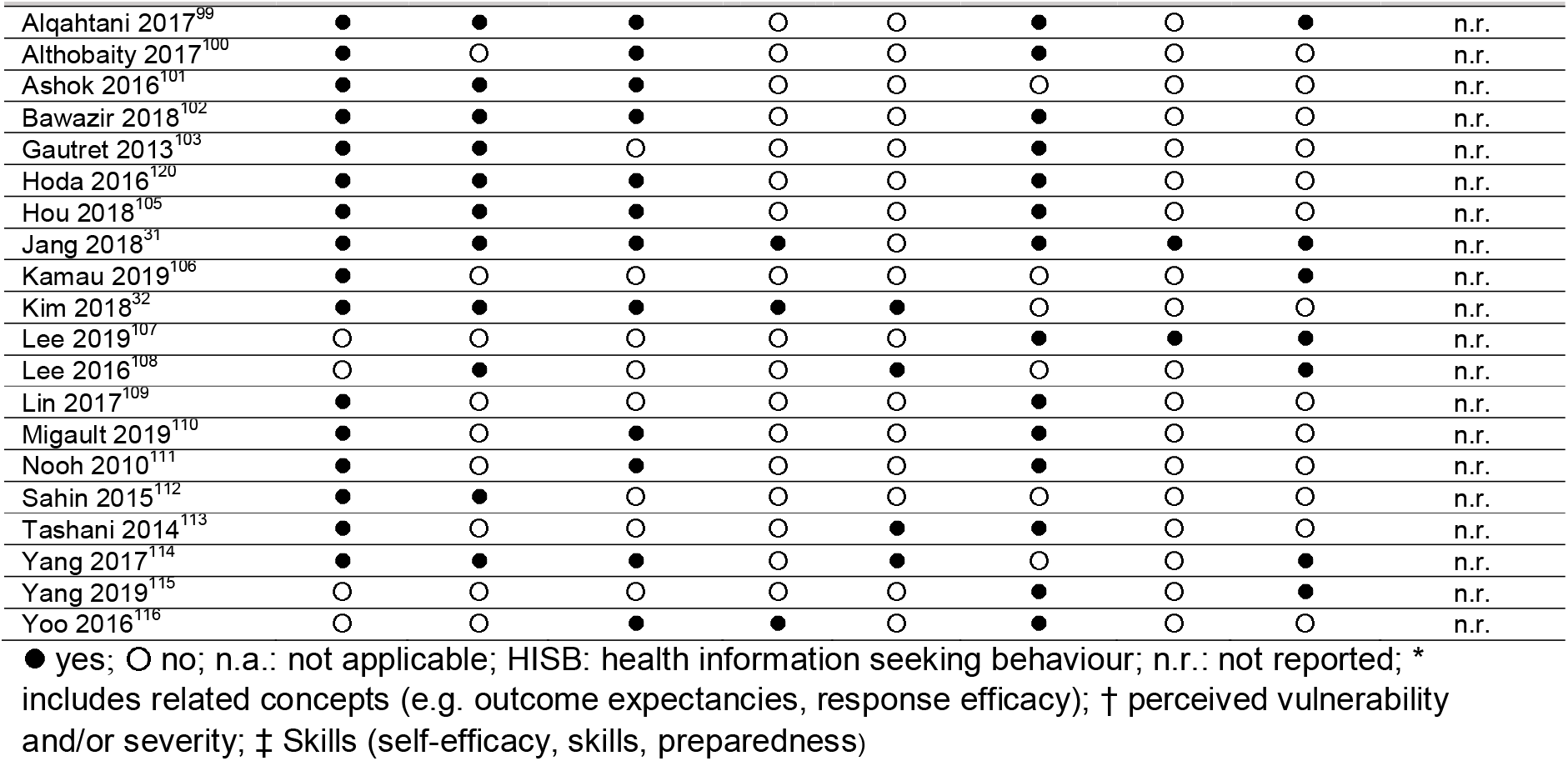

